# Under what circumstances could vaccination offset the harm from a more transmissible variant of SARS-COV-2 in NYC? Trade-offs regarding prioritization and speed of vaccination

**DOI:** 10.1101/2021.01.29.21250710

**Authors:** Hae-Young Kim, Anna Bershteyn, Jessica B. McGillen, R. Scott Braithwaite

## Abstract

**Introduction:** New York City (NYC) was a global epicenter of COVID-19. Vaccines against COVID-19 became available in December 2020 with limited supply, resulting in the need for policies regarding prioritization. The next month, SARS-CoV-2 variants were detected that were more transmissible but still vaccine-susceptible, raising scrutiny of these policies. In particular, prioritization of higher-risk people could prevent more deaths per dose of vaccine administered but could also delay herd immunity if the prioritization introduced bottlenecks that lowered vaccination speed (the number of doses that could be delivered per day). We used mathematical modeling to examine the trade-off between prioritization and the vaccination speed.

**Methods:** A stochastic, discrete-time susceptible-exposed-infected-recovered (SEIR) model with age- and comorbidity-adjusted COVID-19 outcomes (infections, hospitalizations, and deaths by July 1, 2021) was used to examine the trade-off between vaccination speed and whether or not vaccination was prioritized to individuals age 65+ and “essential workers,” defined as including first responders and healthcare, transit, education, and public safety workers. The model was calibrated to COVID-19 hospital admissions, hospital census, ICU census, and deaths in NYC. Vaccination speed was assumed to be 10,000 doses per day starting December 15^th^, 2020 targeting healthcare workers and nursing home populations, and to subsequently expand at alternative starting times and speeds. We compared COVID-outcomes across alternative expansion starting times (January 15^th^, January 21^st^, or February 1^st^) and speeds (20,000, 30,000, 50,000, 100,000, 150,000, or 200,000 doses per day for the first dose), as well as alternative prioritization options (“yes” versus “no” prioritization of essential workers and people age 65+). Model projections were produced with and without considering the emergence of a SARS-COV-2 variant with 56% greater transmissibility over January and February, 2021.

**Results:** In the absence of a COVID-19 vaccine, the emergence of the more transmissible variant would triple the peak in infections, hospitalizations, and deaths and more than double cumulative infections, hospitalizations, and deaths. To offset the harm from the more transmissible variant would require reaching a vaccination speed of at least 100,000 doses per day by January 15^th^ or 150,000 per day by January 21^st^. Prioritizing people ages 65+ and essential workers increased the number of lives saved per vaccine dose delivered: with the emergence of a more transmissible variant, 8,000 deaths could be averted by delivering 115,000 doses per day without prioritization or 71,000 doses per day with prioritization. If prioritization were to cause a bottleneck in vaccination speed, more lives would be saved with prioritization only if the bottleneck reduced vaccination speed by less than one-third of the maximum vaccine delivery capacity. These trade-offs between vaccination speed and prioritization were robust over a wide range of delivery capacity.

**Conclusions:** The emergence of a more transmissible variant of SARS-CoV-2 has the potential to triple the 2021 epidemic peak and more than double the 2021 COVID-19 burden in NYC. Vaccination could only offset the harm of the more transmissible variant if high speed were achieved in mid-to late January. Prioritization of COVID-19 vaccines to higher-risk populations saves more lives only if it does not create an excessive vaccine delivery bottleneck.

## Introduction

In the winter of 2020, New York City (NYC) faced a coalescence of two major developments in the SARS-CoV-2 pandemic: the availability of COVID-19 vaccines and the identification of new variants of SARS-CoV-2. COVID-19 vaccines became available in mid-December 2020, initially with limited supplies targeted to healthcare workers and nursing home populations. Eligibility expanded in January 2021 to include age 65 or older and essential workers, defined as first responders and healthcare, transit, education, and public safety workers.

SARS-CoV-2 variants of concern were reported in Europe, Africa, and South America in January 2021. The lineage B.1.1.7 was first detected in the United Kingdom, where it grew from a rare variant to the dominant circulating variant over a six-week period. The first two NYC cases of B.1.1.7 were confirmed in people who had been infectious at the end of December. Based on its rate of growth in the UK, B.1.1.7 is estimated to be more infectious than previously dominant variants by a factor of 56% (95% CI -8% to +128%).^1^

The emergence of a more transmissible variant could have important implications for the role of vaccination in combatting COVID-19 in NYC. Vaccination has the potential save more lives in the context of a more transmissible variant, compared to if the new variant were never to emerge. Therefore, the first hypothesis in our analysis was the new variant may increase the importance of vaccination speed, i.e., the number of vaccine doses delivered per day, relative to the importance of ensuring vaccines are delivered to priority populations before being offered to others.

The potential trade-off of vaccination speed and prioritization is especially important because concerns have been raised in NYC and elsewhere about bottlenecks in vaccine delivery, as evidenced by the low proportion of ever-stocked vaccines that had been administered to patients in December and early January.^2^ Some hypothesized that removing prioritization guidelines for high-risk populations could reduce these bottlenecks and increase vaccination speed.^3^ Our second hypothesis is that (i) there is a trade-off between prioritization and vaccination speed, and (ii) this trade-off varies based on whether or not a more transmissible variant emerges. Prioritization bottlenecks may be more harmful in the context of a more transmissible variant because of lost opportunities to slow epidemic growth.

To test these hypotheses, we used a mathematical model to compare the impact of vaccination at different speeds with and without emergence of a more transmissible variant in NYC. We assessed the maximum prioritization bottleneck under which prioritization of high-risk groups would still avert more deaths than vaccination without prioritization.

## Methods

### Mathematical model

We augmented a stochastic, discrete-time susceptible-exposed-infected-recovered (SEIR) model of SARS-CoV-2 transmission^4^ with age- and comorbidity-adjusted COVID-19 outcomes (symptoms, hospitalization, ICU admission, and death) using data on NYC’s distribution of age and chronic conditions from New York Behavioral Risk Factor Surveillance System Data in 2017,^5^ the 2013-2014 New York City Health and Nutrition Examination Survey (NYC HANES),^6^ and the US census.^7^ We calibrated the model to COVID-19 in NYC using data on daily COVID-19 hospital admissions, hospital census, ICU census, and deaths from the NYC Department of Health and Mental Hygiene (DoHMH) and Hospital Emergency Response Data System (HERDS). Diminution in R_0_ due to mask-wearing and social distancing was benchmarked based on the observed growth rate in detected cases in early January 2021 after adjustment for percent positivity. We extrapolated R_0_ forward, with or without a linear increase in R_0_ over the period from January 15^th^ to February 28^th^ up to a 56% increase representing the emergence of a more transmissible variant.

### Assumptions about vaccination

Vaccination was assumed to reach 95% efficacy starting 11 days after the first dose and last at least 6.5 months (from the earliest dose delivered on December 15^th^, 2020, until the end of the period of analysis on July 1^st^, 2021. For vaccines requiring multiple doses, the vaccination speeds reported are for the first dose only. We assumed no waning of immunity from vaccination or natural infection. Willingness to be vaccinated was assumed to be 90% for healthcare workers (HCW) and 70% for non-HCW, with lower rates explored in sensitivity analysis.

In the model, we assumed three priority groups: (1) HCW and nursing home residents; (2) essential workers and people aged 65+ years; and (3) all other people. These priority groups are a simplification of the four-phase rollout-plan announced by the Center for Disease Control (CDC) with two of the phases collapsed: (Phase 1a) HCW and nursing home residents; Phase 1b) essential workers; Phase 1c) people aged 65+ years and people aged 16-64 years with underlying medical conditions; and Phase 2) all other people.)

### Model scenarios

Vaccine delivery rates were assumed to be 10,000 doses per day starting December 15^th^, 2020 targeting healthcare workers and nursing home populations. Starting on January 15^th^, January 21^st^, or February 1^st^, 2021, vaccination rates were assumed to increase to 20,000, 30,000, 50,000, 100,000, 120,000, 150,000, or 200,000 doses per day for the first dose and to either prioritize residents age 65+ and essential workers as defined by Cybersecurity & Infrastructure Security Agency (CISA),^8^ or to be applied homogenously without prioritization.

### Evaluation of hypotheses

To evaluate our hypotheses, we compared the number of infections, hospitalizations, and deaths projected to occur by July 1, 2021 in each scenario. To test our first hypothesis, the model was run over a range of vaccination speeds to determine infections and averted deaths for each vaccination speed with and without the emergence of a more transmissible variant. To test our second hypothesis, we estimated the maximum prioritization bottleneck that would avert more deaths compared to vaccination without prioritization. We compared the trade-off for simulations run with and without the emergence of a more transmissible variant in order to determine how the variant may change the trade-off.

## Results

The emergence of a variant with 56% greater transmissibility would increase by 3-fold the peak in daily COVID-19 infections, hospitalizations, and deaths in NYC in the absence of vaccination (Figure 1a-e) and would add 7,947 COVID-19 deaths in NYC between December 15^th^, 2020 and July 1, 2021 (Figure 1f-g) – doubling the COVID-19 death toll over this period.

**Figure 1.**
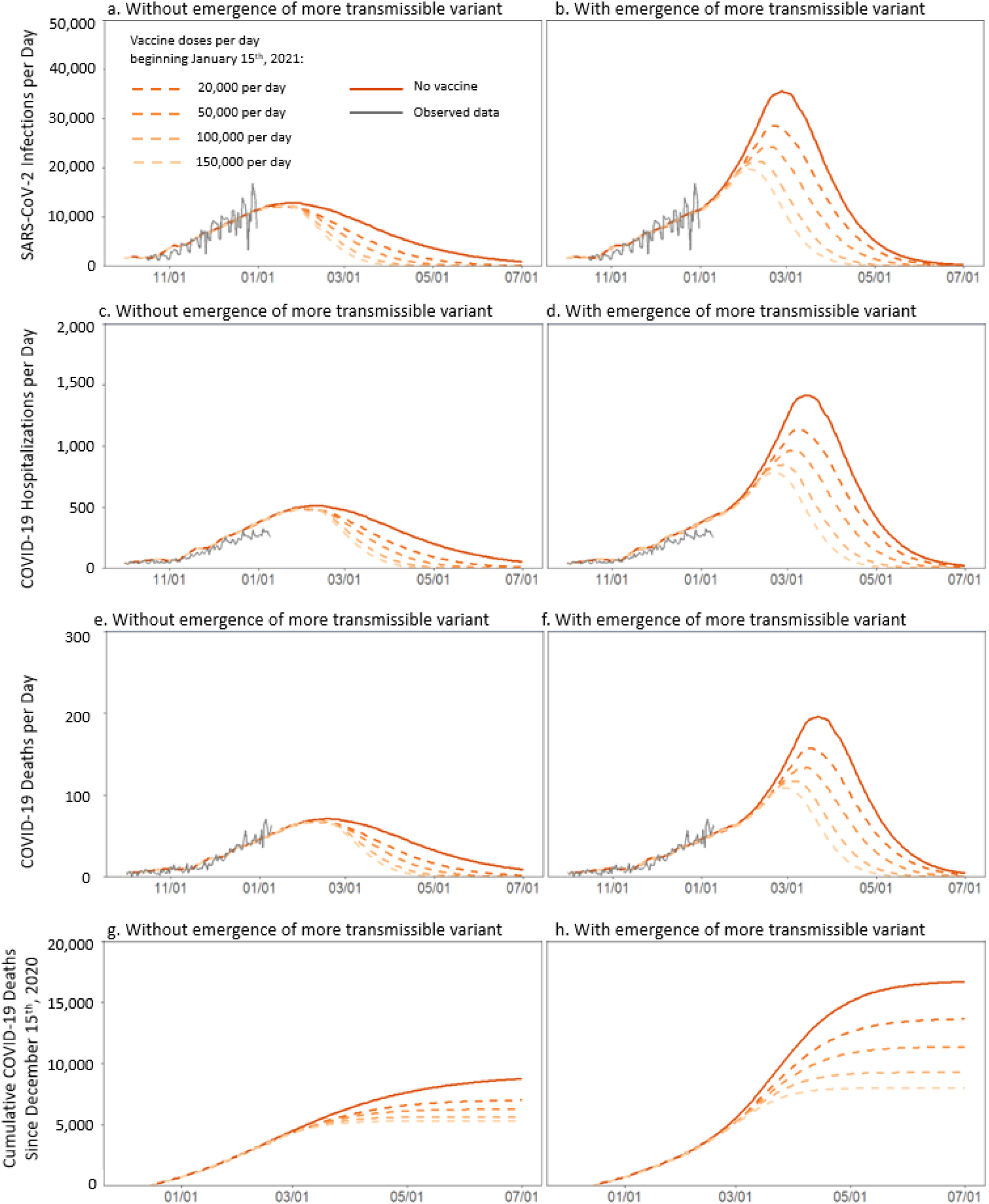
Daily new SARS-CoV-2 infections (a,b), hospitalizations (c,d), and deaths (e,f) as well as cumulative deaths since December 15^th^, 2020 (g.h) in NYC when achieving different vaccination speeds by January 15^th^. Results are computed using model scenarios without (a,c,e,g) and with (b,d,f,h) the emergence of a more transmissible SARS-CoV-2 variant, assumed to gradually increase in prevalence over the period of January 1^st^ through February 28^th^, 2021 with a 56% increase in average SARS-CoV-2 transmissibility by the end of this period. For vaccines requiring multiple doses, the vaccination speeds reported are for the first dose only.

### Impact of vaccination on the harm from a more transmissible variant

The roll-out of vaccination concurrently with emergence of a more transmissible variant could substantially reduce the infections, hospitalizations, and deaths caused by the variant (Figure 1), particularly if the increase in vaccination speed is achieved promptly (by January 15). The slowest-examined vaccination speed of 20,000 doses per day lowers deaths added by the variant from 7,947 to 6,649. The highest-examined vaccination speed of 200,000 doses per day lowers deaths added by the variant further to 2,351, fewer than the deaths that would be averted by similarly-paced vaccination in the absence of the new variant (3,596). Any vaccination speed of 120,000 or more doses per day could offset the harm of the more transmissible variant.

In scenarios where an increase in vaccination speed was not achieved by January 15^th^, but was achieved at a later date (Figure 2b), the benefits of vaccination were smaller overall and required greater speeds to offset the harm of the more transmissible variant. The later the increase in vaccination speed, the smaller the benefit of achieving a given speed. Delivering 150,000 doses per day by January 21^st^ would be required to offset the harm of a more transmissible variant, compared to 120,000 doses by January 15^th^. By February 1^st^, the highest-examined speed of 200,000 doses per day would not be sufficient to offset the harm of the more transmissible variant.

**Figure 2.**
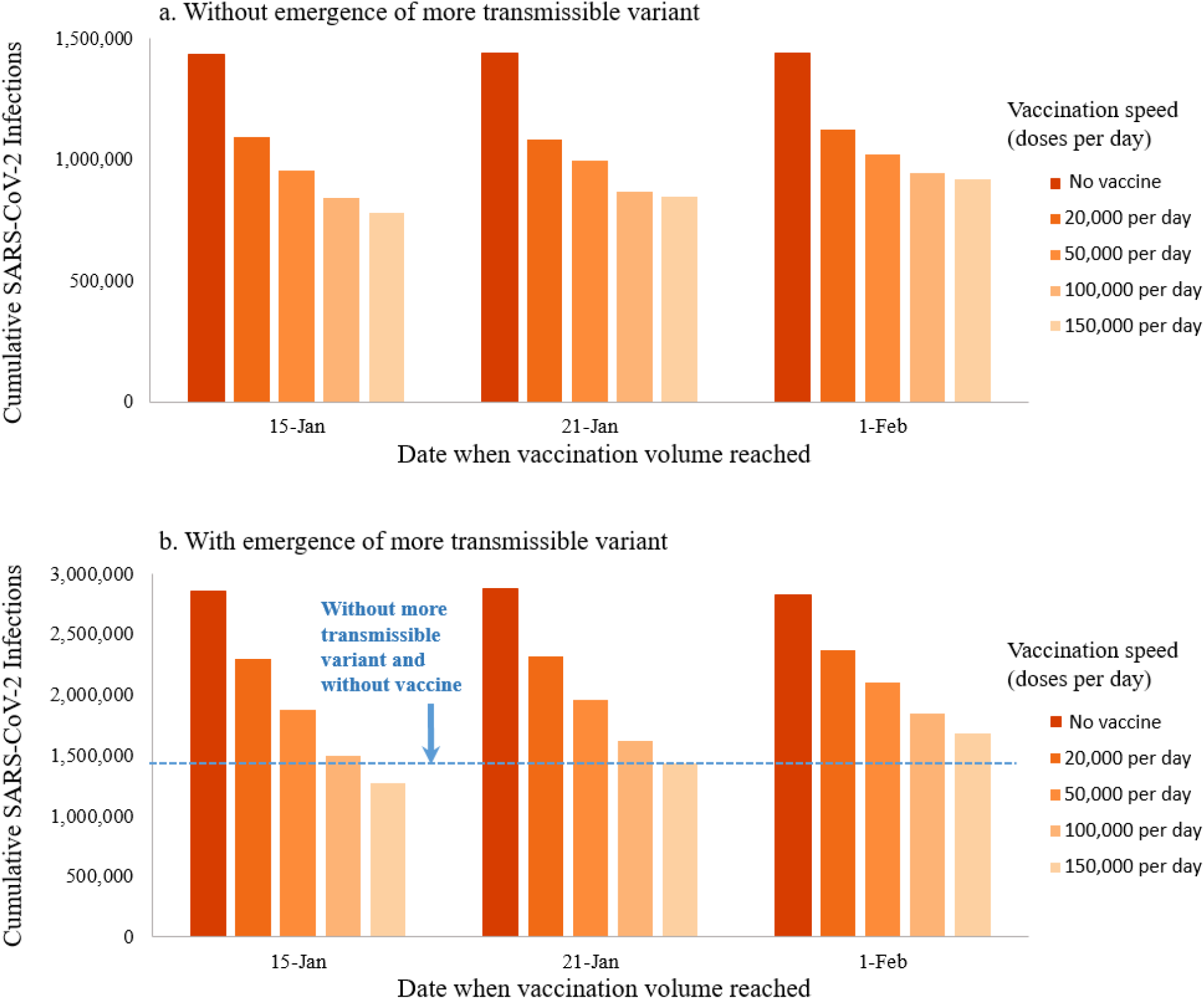
Cumulative SARS-CoV-2 infections over the period Dec 15^th,^ 2020 to July 1^st^, 2021 when achieving different vaccination speeds by January 15^th^, January 21^st^, or February 1^st^, 2021. Results are computed using model scenarios without (a) and with (b) the emergence of a more transmissible SARS-CoV-2 variant, assumed to gradually increase in prevalence over the period of January 1^st^ through February 28^th^, 2021 with a 56% increase in average SARS-CoV-2 transmissibility by the end of this period. For vaccines requiring multiple doses, the vaccination speeds reported are for the first dose only.

### Trade-off between vaccination speed and prioritization to high-risk groups

Prioritization averts more deaths per vaccine dose delivered, as compared to vaccinating uniformly irrespective of priority group. Saving 8,000 lives – nearly one in 1,000 New Yorkers – requires 115,000 doses per day without prioritization but only 71,000 doses per day with prioritization in the context of a more transmissible variant (Table 1).

**Table 1.** Vaccination speed (doses per day beginning on January 15^th^, 2020) needed to avert different numbers of deaths in NYC over the period Dec 15^th,^ 2020 to July 1^st^, 2021. For vaccines requiring multiple doses, the vaccination speeds reported are for the first dose only.

| Deaths<br>averted | Without new variant |  | With new variant |  |
| --- | --- | --- | --- | --- |
|  | No prioritization | Prioritization** | No prioritization | Prioritization** |
| 2,000 | 29,000 | 18,000 | 11,000 | 8,000 |
| 2,500 | 50,000 | 31,000 | 15,000 | 10,000 |
| 3,000 | 83,000 | 50,000 | 19,000 | 13,000 |
| 3,500 | 163,000 | 81,000 | 27,000 | 16,000 |
| 4,000 | >200k | 175,000 | 34,000 | 20,000 |
| 6,000 | >200k | >200k | 61,000 | 40,000 |
| 8,000 | >200k | >200k | 115,000 | 71,000 |

However, if prioritization reduces vaccination speed by causing an implementation bottleneck, the benefits are substantially reduced, and this reduction is proportionately greater in the presence of a more transmissible variant. Without a more transmissible variant, prioritization would only avert deaths if any resulting bottlenecks slowed vaccination speed by less than 50% (Figure 3a). slowed vaccination speed by no more than 33% (Figure 3b).

**Figure 3.**
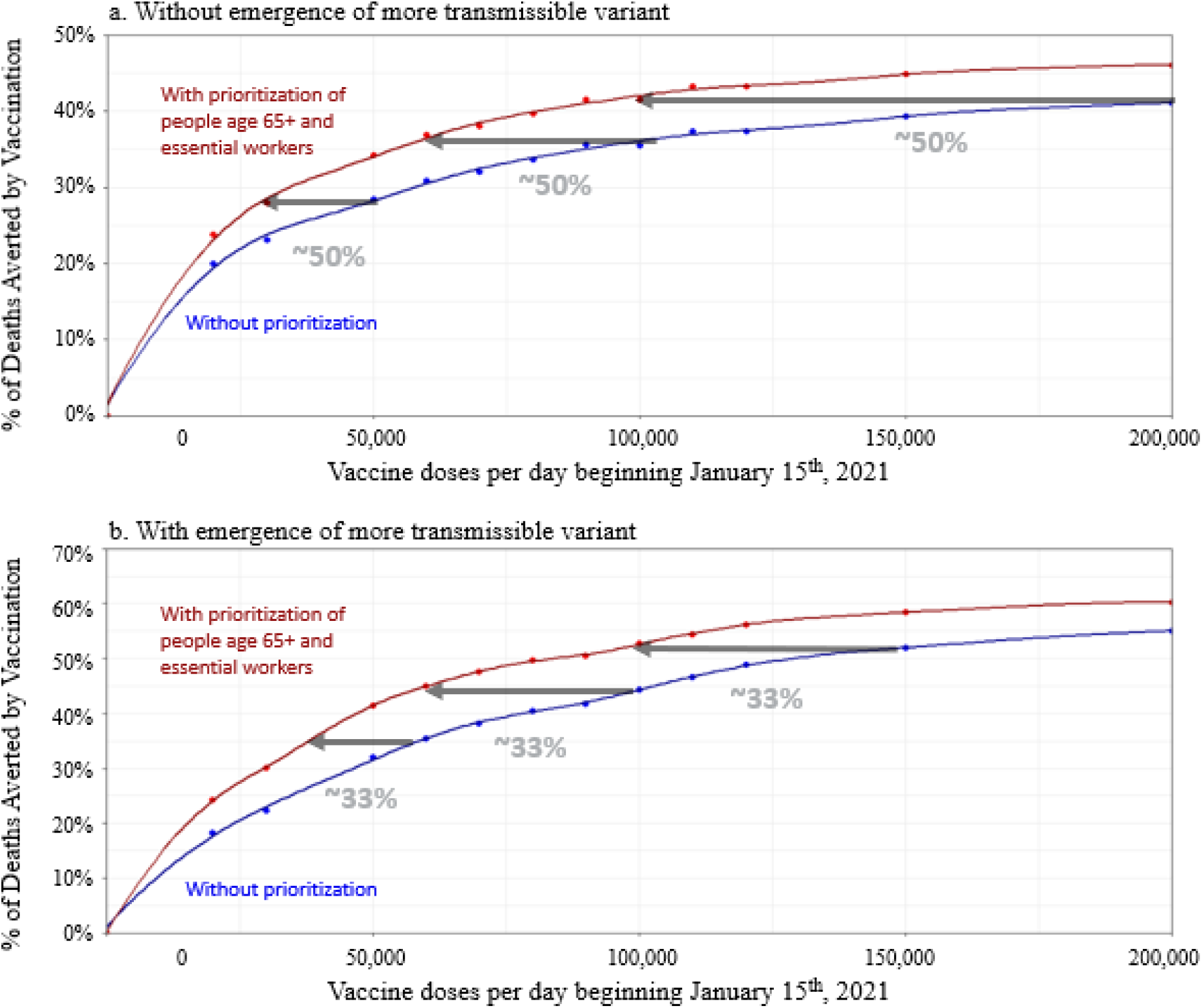
Percent of COVID-19 deaths averted over the period Dec 15^th,^ 2020 to July 1^st^, 2021 by achieving different vaccination speeds by January 15^th^, 2021, compared to no vaccination beyond January 15^th^, 2021. Results are computed using model scenarios without (a) and with (b) the emergence of a more transmissible SARS-CoV-2 variant, assumed to gradually increase in prevalence over the period of January 1^st^ through February 28^th^, 2021 with a 56% increase in average SARS-CoV-2 transmissibility by the end of this period. Red lines show deaths averted by vaccination with prioritization of individuals age 65+ and essential workers. Blue lines show deaths averted by vaccination without prioritization. Gray arrows demonstrate the prioritization bottleneck above which prioritization would no longer be favorable in terms of averting the most deaths. Dots show results of model runs, while lines show the cubic interpolation that was used to estimate the impact of vaccination speeds in-between the values directly evaluated in the model.

## Discussion

The emergence of a more transmissible SARS-CoV-2 variant in NYC would greatly exacerbate the COVID-19 epidemic. Our modeling suggests that in the absence of a vaccine and without substantial changes in behaviors and policies related to the spread of SARS-CoV-2, the variant would triple the 2021 epidemic peak and more than double the 2021 COVID-19 burden.

Harms from the more transmissible variant can be mitigated and potentially abrogated by prompt and speedy vaccination. However, offsetting the added harm of the variant would require vaccination speeds several-fold higher than those achieved by mid-January in NYC. By February 1^st^, it would be too late to offset the harm of the more transmissible variant even if vaccination could reach 200,000 doses per day.

The emergence of a more transmissible variant changes the trade-off between speed and prioritization. Without the more transmissible variant, prioritizing individuals age 65+ and essential workers could avert more deaths even if prioritization-induced bottlenecks reduced vaccination speed by up to half, but with the variant, prioritizing could only avert more deaths if resulting bottlenecks reduced vaccination by up to one-third.

Our analysis has several important limitations. The model did not stratify SARS-CoV-2 transmission patterns according to age, occupation, or neighborhood structure, nor did it incorporate the tendency for a relatively small proportion of individuals to produce a disproportionately large number of secondary cases – a phenomenon known as superspreading. We assumed, based on preliminary evidence, that COVID-19 vaccines would be effective not only against COVID-19 disease but also against either SARS-CoV-2 infection or onward transmission. We further assumed that children would be included in the vaccination program even though, at the time of writing, there are no approved pediatric COVID-19 vaccines.

Assumptions about the emerging SARS-CoV-2 variant were based on the B.1.1.7 lineage, which at the time of writing was the only variant of concern to be confirmed in New York City. The assumed increase in transmissibility was based on preliminary epidemiological data from the United Kingdom which at the time of writing had not been peer-reviewed.^1^ Our simulations also did not consider differences in pathogenicity among different variants, nor immune evasion by new variants. Evidence on these topics was still emerging at the time of writing and may require updated assumptions.

These limitations may bias our results in different ways. If prioritized occupations such as healthcare, education, and transit contribute more to SARS-CoV-2 transmission than other population groups, then our results may be biased in favor of vaccination speed rather than prioritization by failing to account for the transmission benefit of prioritizing essential workers. On the other hand, if even more transmissible variants than the B.1.1.7 lineage are destined to arrive in NYC, or if new variants are capable of re-infecting recovered individuals by evading naturally-acquired immunity, then NYC may expect to see a larger number of future infections than were projected by our model, and our results may be biased in favor of prioritization rather than vaccination speed.

## Conclusions

Our modeling emphasizes the urgency of rapid, high-speed COVID-19 vaccination in the context of the emergence of a transmissible variant of SARS-CoV-2 in NYC. The emergence of a more transmissible variant of SARS-CoV-2 has the potential to triple the 2021 epidemic peak and more than double the 2021 COVID-19 burden in NYC. Vaccination could only offset the harm of the more transmissible variant if high vaccination speed were achieved in mid-to late January. Prioritization of COVID-19 vaccines to higher-risk populations saves more lives only if it does not create an excessive vaccine delivery bottleneck.

## Data Availability

The data on NYC COVID-19 epidemic trends is publicly available by the New York City Department of Health and Mental Hygiene.

https://github.com/nychealth/coronavirus-data

## Acknowledgements

This work was supported by the New York City Department of Health and Mental Hygiene.

